# Cannabis use and Cancer: Dissecting genetic causality for site-specific risks through two-sample Mendelian Randomization

**DOI:** 10.64898/2026.08.12.26360176

**Authors:** Elena Lukhere, Baxter Kachingwe, Wakisa Kipandula, Noel Chiphangwi, Mwiza Gideon Singini, Abram Bunya Kamiza

## Abstract

**Background:** The prevalence of cannabis use is increasing at an alarming rate owing to its legalization and decriminalization in some countries. Epidemiological evidence on the association between cannabis use and cancer is inconsistent and conflicting. Herein, we performed two-sample Mendelian randomization (MR) to investigate whether cannabis use is causally associated with site-specific cancers in individuals of European ancestry.

**Methods:** We identified 22 independent genetic variants strongly associated with cannabis use (p-value < 5 x 10^-8^) in a large meta-analysis of genome-wide association studies of individuals of European ancestry. Genome-wide association summary-level data on site-specific cancers were obtained from individuals of European ancestry in FinnGen, Finland. MR analyses were performed using the inverse-variance weighted (IVW) and multivariable method. Sensitivity analyses were performed using the simple median, weighted median, MR-Egger, and MR pleiotropy residual sum and outlier methods.

**Results:** Our multivariable IVW analyses adjusted for cigarette smoking found that genetic liability to cannabis use was causally associated with esophageal cancer (odds ratio [OR] =1.74, 95% confidence interval [CI] =1.29-2.15, p-value =0.013) and lung cancer (OR=1.35, 95% CI = 1.13-1.58, p-value =0.009). However, genetic liability to cannabis use exerted a protective effect against pancreatic cancer (OR=0.77, 95% CI =0.57-0.91, pvalue=0.032) in individuals of European ancestry in the FinnGen. Our sensitivity analyses found no evidence of horizontal pleiotropy between cannabis use and site-specific cancers.

**Conclusion:** We found that genetic liability to cannabis use was associated with esophageal, lung, and pancreatic cancers in individuals of European ancestry.

## Introduction

With the legalization and decriminalization of cannabis in some countries, the number of cannabis users has increased rapidly ^1^. In the US, the prevalence of cannabis use is more than 55% among the adult population ^2,3^. In the UK, approximately 30% of the population uses cannabis regularly ^4^. Cannabis, also known as marijuana, contains more than 600 chemical compounds, of which 60 are cannabinoids ^5^. Cannabis use, especially through smoking, has been associated with adverse health outcomes, including bronchitis, emphysema, psychosis, schizophrenia, and cardiovascular diseases^6–8^. Furthermore, evidence indicates that cannabis use is associated with site-specific cancers ^9^.

However, evidence from epidemiological studies investigating the association between cannabis use and cancer is inconsistent and conflicting ^10–19^. Some evidence indicates that cannabis use is associated with an increased risk of various cancers, including lung ^10,11^, prostate ^12^, testicular ^13–15^, and head and neck cancers ^16^. However, other evidence suggests that cannabis use is not associated with breast, cervical, colorectal, lung, or head and neck cancer ^12,17–19^. Further evidence has shown that cannabis use is inversely associated with bladder, head and neck cancers ^9,20,21^. The inconsistent results reported thus far may be due to response bias, as cannabis use is still illegal in most countries, misclassification of cannabis users, recall bias, ascertainment bias, and failure to adjust for confounders that are strongly associated with cannabis use, including cigarette smoking. Moreover, most studies performed thus far have been observational studies that are prone to confounding and reverse causation; hence, they cannot be used to infer causality.

Nevertheless, a Mendelian randomization (MR) study was performed to assess whether cannabis use is causally associated with site-specific cancers in individuals of European ancestry ^22^. Huang et al 2023 found cannabis use to be significantly associated with cancer of the cervix and reported suggestive significance for cancer of the breast and laryngeal ^22^. This study requires further validation as it was limited by the lack of instrumental variables that were not strongly associated with cannabis use, which may have decreased the statistical power to detect the association between cannabis use and site-specific cancers. Moreover, cigarette smoking, which is strongly associated with cannabis use, was not corrected for in this study..

Given these limitations, we conducted an MR study to investigate whether cannabis use is associated with site-specific cancers using instrumental variables that are strongly associated (p-value <5 x10^-08^) with the exposure (cannabis use) and also adjusting for potential confounders of cigarette smoking using multivariable MR approaches. MR studies improve causal inference using genetic variants as instrumental variables for the association between exposure and outcome variables ^23^. MR is not susceptible to reverse causation, as genetic variants are fixed at birth and are not affected by unmeasured factors, which usually confound epidemiological studies ^24^. This makes MR an ideal approach for investigating the association between cannabis use and site-specific cancers. Herein, we performed two-sample MR to investigate whether cannabis use is causally associated with site-specific cancers in individuals of European ancestry. The overview of the study is shown in **Fig. 1**

**Fig. 1.**
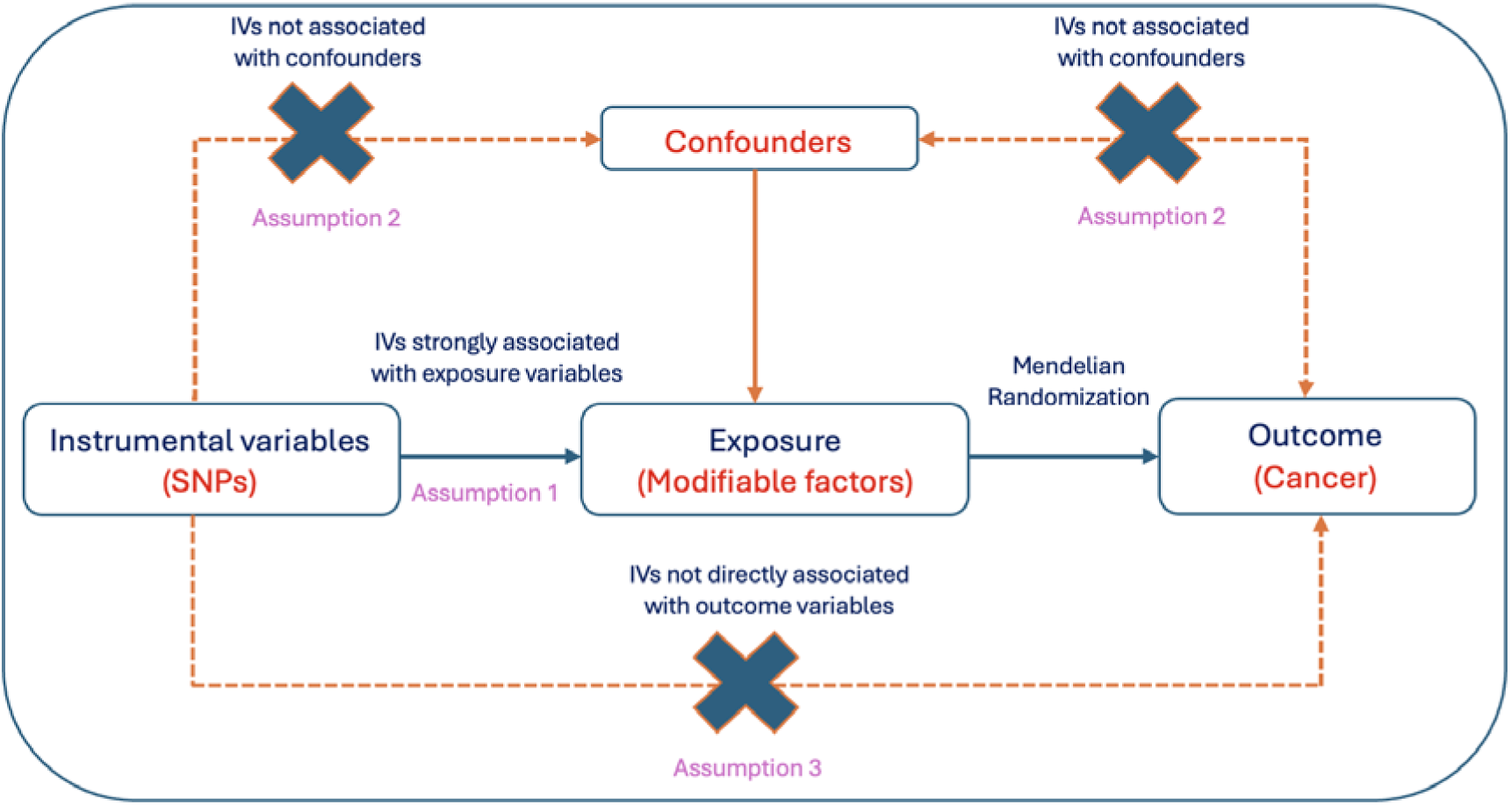
Overview of the study showing the assumption of Mendelian randomization.

## Methods

### Exposure data

Genetic variants strongly associated with cannabis use were selected from a large meta-analysis of genome-wide association studies (GWASs) of individuals of European ancestry^25^. Cannabis use was defined based on participant cannabis dependence or abuse according to the International Classification of Disease (ICD) 9 and 10 codes. In individuals of European ancestry, the meta-analysis included 42,281 cannabis dependencers or abusers and 843,744 controls^25^. We found 1,274 single nucleotide polymorphisms (SNPs) strongly associated with cannabis use at a genome-wide significant level (p-value < 5 x 10^-8^) ^25^. We then applied distance clumping for all significant SNPs (±500kb) and found 23 lead SNPs that were strongly associated with cannabis use. Of these SNPs, one was palindromic and was subsequently removed for further analysis (Fig.2). Ultimately, we used 22 independent genetic variants as the instrumental variables. To avoid horizontal pleiotropy, we used PhenoScanner ^26^ to identify and remove SNPs directly associated with site-specific cancers. Interestingly, our instrumental variables were not directly associated with any site-specific cancers; the minimum F-statistic for our instrumental variables was 58.2 (**Table. S1**), indicating that our instrumental variables were strongly associated with cannabis use.

**Fig. 2.**
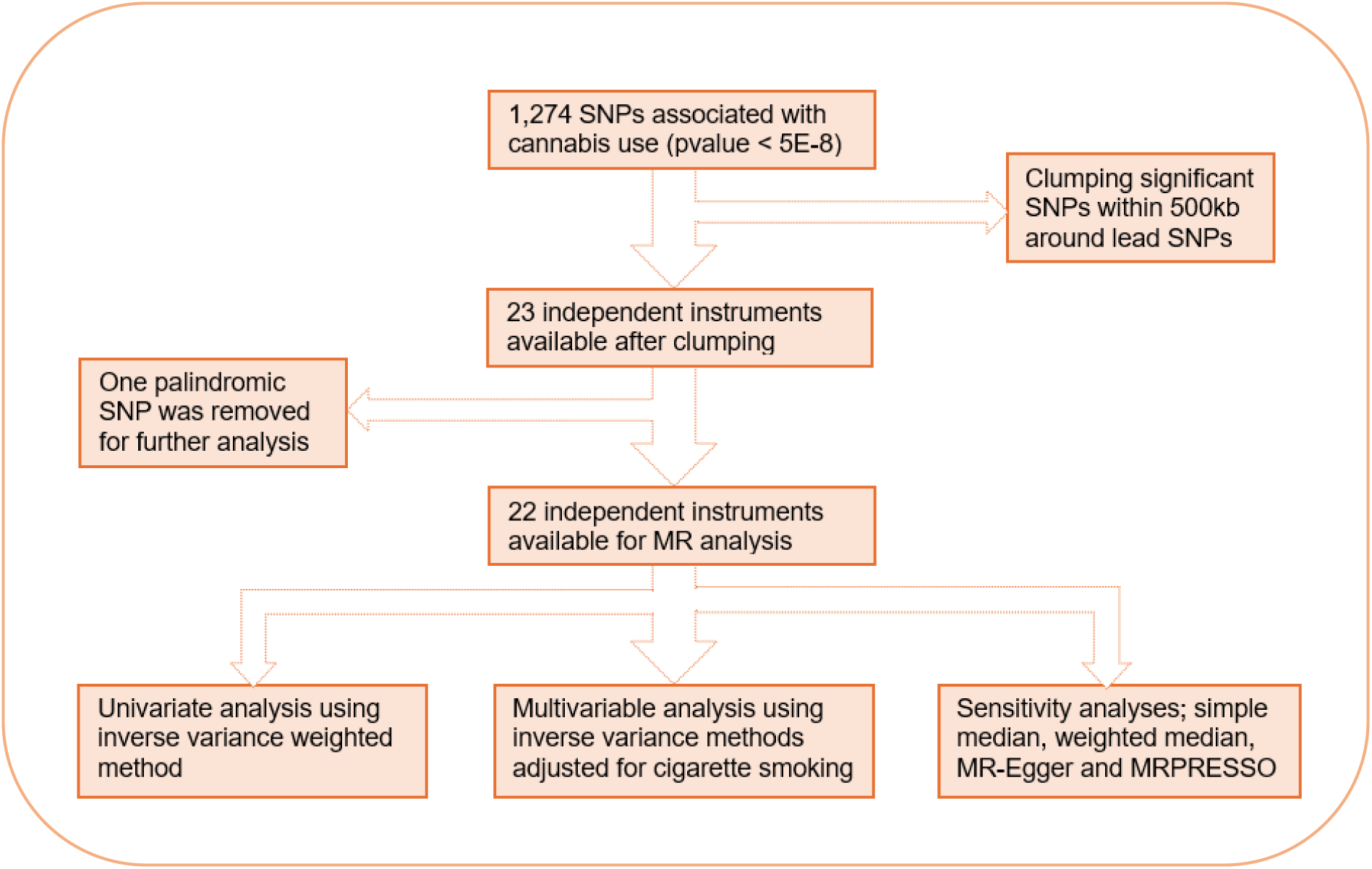
Flowchart of instrumental variable section and statistical approaches used in Mendelian randomization.

### Outcome data

The GWAS summary statistics results for site-specific cancers, including cancer of the biliary, bladder, brain, breast, cervix, colon, esophagus, gastric, head and neck, kidney,lung, oral, ovary, pancreas prostate, rectum, skin, testis, thyroid, and urinary were obtained from individuals of European ancestry in the FinnGen, Finland ^27^. Finngen was set up with the aim of providing new insights into disease genetics. The project combined imputed genotyped data from Finnish biobanks and electronic health record from Finnish registries. Finngen has so far collected samples from 412,000 individuals across Finland. Our two-sample MR study takes advantage of publicly available GWAS summary statistics results of various site-specific cancers obtained from https://www.finngen.fi/en

### Ethics statement

This study used publicly available GWAS summary statistics from the largest multi-ancestry meta-analysis of cannabis use disorder ^25^ and FinnGen. The original studies obtained informed consent from the participants and were conducted in accordance with protocols approved by the relevant institutional review boards and ethics committees. No additional ethical approvals were required.

### Statistical analysis

The inverse-variance weighted (IVW) method was the main MR method used in the absence of directional pleiotropy and heterogeneity ^28^. The heterogeneity between the exposure and outcome data was investigated and estimated using Cochran’s Q and I2 statistics ^29^. Sensitivity analyses were performed to detect the presence of pleiotropy^28^. Heterogeneity between exposure and outcome was determined if Cochran’s Q statistics were statistically significant at a p-value < 0.05; consequently, we used a random-effects IVW method ^30^. The MR-Egger test was used to detect the presence of horizontal pleiotropy. When the MR-Egger intercept deviated from zero or its p-value was statistically significant at p-value < 0.05, it indicated the presence of horizontal pleiotropy ^31^, and MR pleiotropy residual sum and outlier (MR-PRESSO) was performed to identify and remove outlier genetic variants^32^. Since cannabis use is strongly associated with cigarette smoking, we performed multivariable MR by including genetic variants strongly associated with cigarette smoking in individuals of European ancestry^33^. For multiple testing, we used the false discovery rate^34^, and the association between cannabis use and cancer was considered statistically significant at a p-value < 0.05. All analyses were conducted using Two-Sample MR and the Mendelian Randomisation ^35^ packages in R version 4.1.2

## Results

### Univariate MR analyses

We identified 22 independent genetic variants that were strongly associated with cannabis use from a large GWAS meta-analysis (TableS.1). Univariate IVW was our main MR method in the absence of heterogeneity and horizontal pleiotropy. We found that genetic liability to cannabis use was causally associated with cervical cancer (odds ratio [OR]= 1.66, 95% confidence interval [CI]= 1.26-2.06, p-value = 0.022), esophageal cancer (OR=1.63, 95% CI = 1.28-1.98, p-value =0.012), and lung cancer (OR = 1.34, 95% CI =1.12-1.56, p-value=0.009, **Fig.3**) in individuals of European ancestry. However, we found that genetic liability to cannabis use exerted a protective effect against pancreatic cancer (OR=0.76, 95%CI =0.54-0.87, p-value=0.018).

**Fig. 3.**
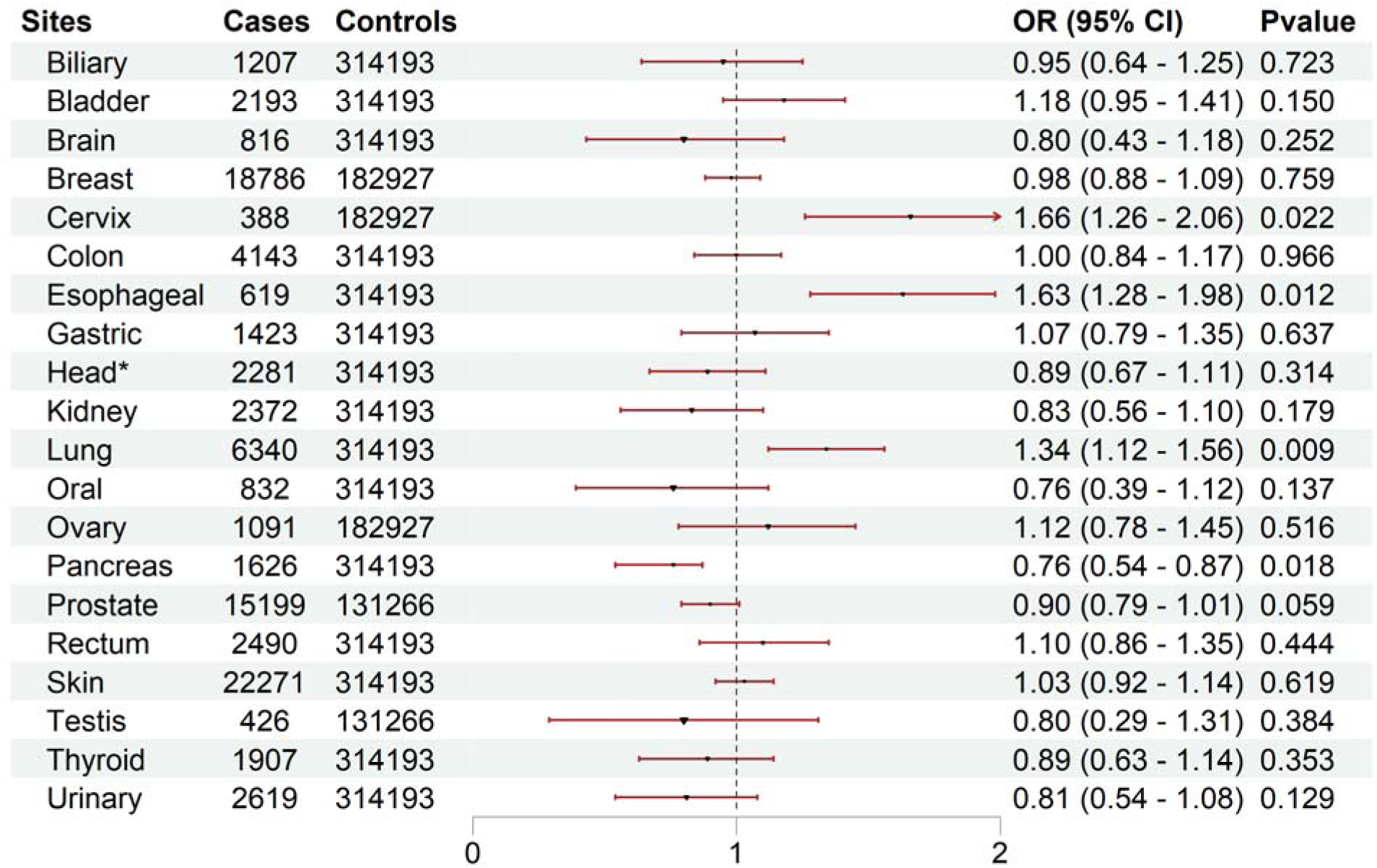
Univariate Mendelian randomization between cannabis use and site-specific cancer in FinnGen, Finland. * Head and neck cancer

### Multivariable MR analyses

We then performed multivariable IVW analyses adjusted for cigarette smoking and found that genetic liability to cannabis use was causally associated with esophageal cancer (OR=1.74, 95% CI=1.29-2.15, p-value =0.013) and lung cancer (OR=1.35, 95% CI = 1.13-1.58, p-value=0.009). However, our multivariable IVW analysis found that genetic liability to cannabis use exerted a protective effect against pancreatic cancer (OR=0.77, 95% CI =0.57-0.91, p-value =0.032) after adjusting for cigarette smoking (**Fig.4**). Although our univariate MR found an association between cannabis use and cervical cancer, our multivariate MR results for cannabis use and cervical cancer were not statistically significant (OR=1.63, 95% CI=0.97-1.98, p-value=0.076) after adjusting for cigarette smoking, suggesting that the results were confounded by cigarette smoking.

**Fig. 4.**
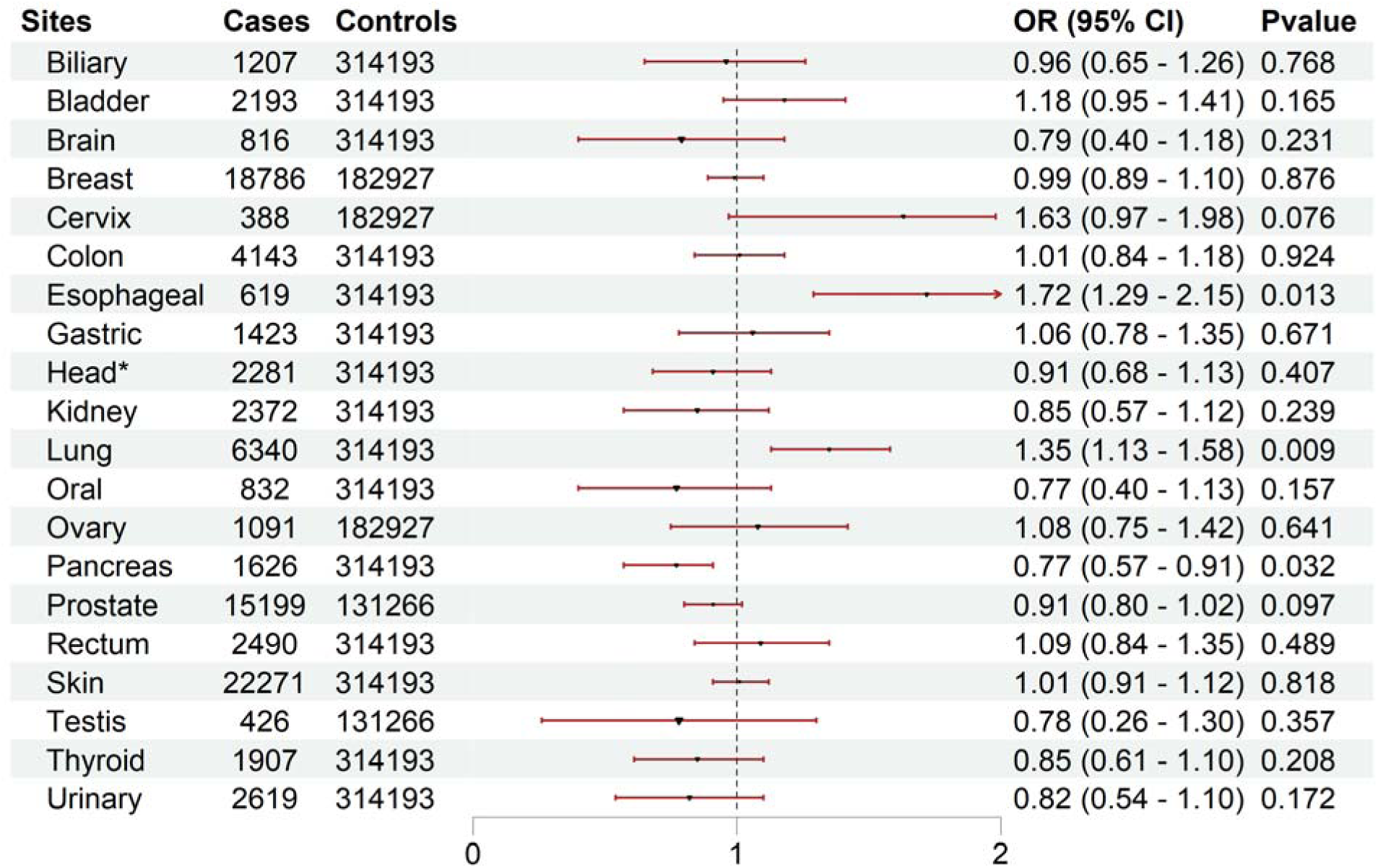
Multivariate mendelian randomization between cannabis use and site-specific cancer in FinnGen, Finland adjusted for cigarette smoking. * cancer of head and neck

### Sensitivity analyses

Sensitivity tests were performed using different MR approaches, including the MR-PRESSO, MR egger, simple median, and weighted median methods. Our sensitivity analysis found no evidence of horizontal pleiotropy for the exposure and outcome, with p-values >0.05 for the MR-Egger regression intercept. Furthermore, we found no evidence of heterogeneity between cannabis use and site-specific cancers. We then used MR-PRESSO to identify and remove any outlier instrumental variables driving the association between cannabis use and cancer. Interestingly, our analysis did not identify any outlier instrumental variables driving the association between cannabis use and cancer, suggesting that our results were statistically robust.

## Discussion

We set out this study to investigate whether genetic liability to cannabis use was causally associated with various site-specific cancers in individuals of European ancestry. Using multivariable MR analysis adjusted for cigarette smoking, we found that genetic liability to cannabis use was causally associated with esophageal and lung cancers. However, genetic liability to cannabis use was associated with a decreased risk of pancreatic cancer in individuals of European ancestry in Finland.

Our MR analysis found that genetic liability to cannabis use was causally associated with lung cancer, corroborating previous analyses ^10,11^. The increased risk of lung cancer among cannabis users may be due to the presence of well-known carcinogens, including nitrosamines, polycyclic aromatic hydrocarbons, benzopyrene, vinyl chlorides, and phenols in cannabis smoke ^7–9^, which may initiate and promote tumorigenesis among cannabis users. Moreover, a previous study indicated that cannabis smoke contains more carcinogens than tobacco smoke ^36^. Furthermore, cannabis smoking behaviours, such as the use of unfiltered cigarettes, deep inhalation, and prolonged breath-holding ^37^, may exacerbate exposure to cytotoxic and mutagenic agents, which may initiate mutagenesis in the lower respiratory tract. Experimental studies have also demonstrated that regular cannabis smokers manifest airway inflammation, in addition to histopathological and molecular changes in the bronchus, indicative of precancerous activity ^38,39^. However, other studies have failed to find any association between cannabis use and lung cancer ^17,18^. The non-significant findings reported by previous studies may be due to the misclassification of cases and small sample sizes, which decreased the statistical power to detect the association between cannabis use and lung cancer. In this analysis, we used instrumental variables that were strongly associated with cannabis use, as defined by cannabis dependence and abuse by ICD 9 and 10 codes.

Our multivariate IVW method adjusted for cigarette smoking found genetic liability to cannabis use to be causally associated with esophageal cancer. The association between cannabis use and esophageal cancer has rarely been investigated previously. Notably, only one observational study has investigated this association and found that cannabis use was not associated with esophageal cancer ^40^. The nonsignificant results reported by Hashibe et al. (2006) may be due to the limited number of long-term cannabis users in their study ^40^, which decreased the statistical power of detecting the association between cannabis use and esophageal cancer. The mechanism linking cannabis use to esophageal cancer has not yet been fully elucidated. However, previous studies have suggested that cannabis may contribute to the development of Barrett’s esophagus and gastroesophageal reflux disease^41,42^, which are the main risk factors for esophageal cancer.

Notably, this is the first study to assess the association between cannabis use and pancreatic cancer. We found that cannabis use reduced the risk of developing pancreatic cancer in individuals of European ancestry after adjusting for cigarette smoking. The endocannabinoid system, through which cannabinoids operate, regulates and controls many bodily functions and is widely distributed throughout the gastrointestinal tract with regional variations and organ-specific actions^43^. Despite having more than 60 active cannabinoid compounds, research on cannabis has stalled over the years because cannabis is still illegal in most countries. Of the 60 active cannabinoid compounds found in cannabis, only four have been extensively studied, including CBD, cannabinol, d-8-THC, and d-9-THC ^5^. The most common of these compounds are d-9-THC and CBD, which inhibit tumorigenesis by modulating important signalling pathways, including the cell cycle, apoptosis, and angiogenesis^44,45^. However, these cannabinoids may also have a carcinogenic effect and promote tumor growth by inducing apoptosis in immune cells as a pathway for immunosuppression ^46^. The decreased risk of pancreatic cancer among cannabis users may be due to the organ-specific action of cannabinoids with antineoplastic properties, including promotion of cell cycle arrest, apoptosis, anti-angiogenesis, inhibition of cellular invasion, and migration ^44,45^, which are crucial for tumor initiation, promotion, and development. These antitumoral activities have been observed in other cancers, including brain, prostate, thyroid, skin, colon and breast cancers ^47^. Nevertheless, these site-specific cancers were not statistically significant in this study.

Our study had several strengths, including the use of the MR approach, which eliminated some unmeasured confounders commonly observed in epidemiological studies. Moreover, we used multiple instrumental variables that were strongly associated with cannabis use in individuals of European ancestry. We adjusted for cigarette smoking, which is strongly associated with cannabis use; therefore, our results were statistically robust. In addition, we used a homogenous population that minimized the heterogeneity commonly observed when individuals of different ancestral populations are used in genetic studies. A limitation of this study is the small sample size for some site-specific cancers, including testicular cancer. Another limitation is that our results might not be generalizable to other ancestral populations, as we only included individuals of European ancestry.

In conclusion, we found that cannabis use is causally associated with lung and esophageal cancers in individuals of European ancestry and decreases the risk of pancreatic cancer. Further research is needed to characterize the pharmacological effects of all chemical compounds found in cannabis and to replicate our analysis in other ancestry populations using genetic variants that are strongly associated with cannabis use.

## Data Availability

Genome-wide association study summary statistics supporting the findings of this study are available at https://www.finngen.fi/en

## Authors’ contributions

Conceptualization: ABK; methodology: EL,ABK; writing-original draft: EL, BK, ABK; writing-review and editing: EL, BK, WK, NC, MGS,ABK.

## Competing interests

The authors declare no conflicts of interest.

## Funding

None

## Supplementary materials

**Table S1.** Instrumental variables ssociated with cannabis use in individuals of European ancestry.

| SNP | CHR | BP | MAF | BETA | SE | P-value | F-statistics |
| --- | --- | --- | --- | --- | --- | --- | --- |
| rs1526480 | 1 | 91209986 | 0.5915 | -0.0519 | 0.0084 | 5.906e-10 | 115.4 |
| rs6690119 | 1 | 73580964 | 0.4433 | 0.0467 | 0.0086 | 4.995e-08 | 95.4 |
| rs7519259 | 1 | 66434743 | 0.5298 | 0.0501 | 0.0083 | 1.83e-09 | 110.9 |
| rs719504 | 2 | 22918025 | 0.3499 | 0.0909 | 0.0163 | 2.533e-08 | 334.3 |
| rs17007864 | 3 | 70876858 | 0.3877 | 0.0522 | 0.0086 | 1.053e-09 | 114.7 |
| rs184064410 | 3 | 43992164 | 0.999 | 0.5732 | 0.1006 | 1.202e-08 | 58.2 |
| rs3774800 | 3 | 49334768 | 0.6581 | -0.0598 | 0.0085 | 1.718e-12 | 142.8 |
| rs6790568 | 3 | 49835450 | 0.84 | 0.0628 | 0.0106 | 2.728e-09 | 94.0 |
| rs726610 | 3 | 85551403 | 0.3489 | -0.0563 | 0.0085 | 4.288e-11 | 127.7 |
| rs7616815 | 3 | 48779715 | 0.334 | -0.0497 | 0.0086 | 7.194e-09 | 97.4 |
| rs159365 | 5 | 60500273 | 0.6004 | 0.0461 | 0.0083 | 3.33e-08 | 90.4 |
| rs9344740 | 6 | 88619412 | 0.3062 | -0.0556 | 0.0091 | 8.344e-10 | 116.5 |
| rs62461183 | 7 | 77716309 | 0.162 | 0.0697 | 0.0113 | 5.863e-10 | 117.0 |
| rs2189010 | 7 | 114119430 | 0.4761 | 0.0481 | 0.0085 | 1.28e-08 | 102.3 |
| rs56372821 | 8 | 27436500 | 0.1551 | -0.0887 | 0.0119 | 7.272e-14 | 183.0 |
| rs10986600 | 9 | 127928735 | 0.3091 | 0.057 | 0.009 | 2.172e-10 | 123.1 |
| rs200595759 | 10 | 118683917 | 0.3234 | -0.131 | 0.0231 | 1.42e-08 | 670.4 |
| rs10835372 | 11 | 28643913 | 0.3688 | 0.0471 | 0.0085 | 3.028e-08 | 91.6 |
| rs34554234 | 11 | 113292326 | 0.3487 | -0.0543 | 0.0091 | 2.198e-09 | 118.8 |
| rs6484345 | 11 | 27996573 | 0.8141 | 0.1147 | 0.0203 | 1.634e-08 | 354.2 |
| rs80030908 | 13 | 55159898 | 0.0189 | 0.171 | 0.0305 | 2.127e-08 | 96.1 |
| rs62051488 | 16 | 72652784 | 0.1093 | -0.0725 | 0.0131 | 2.976e-08 | 90.7 |

SNPs; single nucleotide polymorphisms, CHR; chromosomes, BP; base pair position, MAF; minor allele frequency, SE; standard error

## Notes

### Competing Interest Statement

The authors have declared no competing interest.

### Author Declarations

This study used publicly available GWAS summary statistics from the largest multi-ancestry meta-analysis of cannabis use disorder and FinnGen. The original studies obtained informed consent from the participants and were conducted in accordance with protocols approved by the relevant institutional review boards and ethics committees. No additional ethical approvals were required.

